# Expert-level localization of fine-grained cardiac structures across the transthoracic echocardiogram

**DOI:** 10.64898/2026.11.10.26367643

**Authors:** Elham Mahmoudi, Nicolo Pecco, Hesham Sheashaa, Nima Baba Ali, Kamal Awad, Fatmaelzahraa Emad Abdelfattah, Sogol Attaripour Esfahani, Ibrahim Omar, Nadera Naquib Bismee, Isabel G. Scalia, Juan Farina, Chadi Ayoub, Reza Arsanjani, Martha Grogan, Garvan Kane, Jae K. Oh, Imon Banerjee, Bradley J. Erickson, Chieh-Ju Chao

## Abstract

Artificial intelligence models for echocardiography interpret studies without identifying the structures that support their outputs. We show that this spatial grounding does not emerge from the pretraining of current vision-language models. General-purpose and medical vision-language models and an open-vocabulary detector recalled 3% or fewer of cardiac structures and scored below a box that never sees the image. EchoLens, a detector trained on 5,912 expert-annotated structures and 4,219 Doppler signals from 136 patients, recalled 87% and localized structures as reliably as a blinded expert reader (box centre within the reference in 95.7% and 95.3% of cases). It covers 21 structures across 11 transthoracic views and 11 spectral Doppler signal types. In external data, two physicians judged 81– 92% of its detections correct across all 11 views, and it identified the Doppler signal type in 83% of frames. EchoLens provides the spatial layer that grounded echocardiographic interpretation requires.

## 2. Introduction

Echocardiography remains the primary non-invasive imaging modality for evaluating cardiac anatomy and function in various clinical settings^1^. Over the past decade, artificial intelligence (AI) has rapidly transformed quantitative echocardiography. Early deep learning models relied primarily on task-specific convolutional neural networks designed for single or few objectives, such as view classification^2^, left ventricular ejection fraction estimation^3^, or automated interpretation of individual findings^4^.

Foundation models and vision-language models (VLMs) trained on large corpora of echocardiographic videos and reports, such as EchoCLIP^5^, PanEcho^6^ and EchoPrime^7^, now perform multi-task interpretation across the transthoracic examination. However, they operate on whole frames, clips or studies, and their outputs are not tied to the cardiac structures that support them, and a recent echocardiography video-language model grounds its outputs in time, by selecting keyframes, but not in space^8^. Post hoc saliency methods such as Grad-CAM^9^, the usual route to regional interpretation, are poorly suited to these multi-task models with complex architectures. In conventional deep learning models where saliency can be computed, it localizes less accurately than human experts or dedicated localization networks^10,11^. Localizing each structure is the prerequisite for such grounding.

In general computer vision, object detection frameworks such as You Only Look Once (YOLO)^12^ and Detection Transformers (DETR)^13^, together with promptable segmentation models such as Segment Anything (SAM)^14^ and open-vocabulary detectors such as Grounding DINO^15^, provide mature tools for spatial localization. However, within echocardiography, object detection has addressed one or a few targets, such as the left ventricle^16^, the valves in apical views^17^, the chambers or substructures of the fetal four-chamber view^18–20^ and septal defects in congenital heart disease^21,22^, and no detection framework spans the standard adult transthoracic examination.

Here we present EchoLens, a detection framework that localizes cardiac structures across the standard views of the adult transthoracic examination, together with spectral Doppler signals. We hypothesized that fine-grained structure localization does not emerge from the scale or medical pretraining of current vision-language models, and that a dedicated detector trained on a modest set of expert-annotated frames can provide it across views and in external data.

## 3. Results

The development dataset comprised 5,912 annotated cardiac structures (21 structures, 54 structure-view classes, 11 views) on 1,035 B-mode frames and 4,219 spectral Doppler signals (11 signal types) on 1,806 frames, from 136 patients (132 with Doppler), divided at the patient level into training, validation and test sets (Table 1; Methods). External evaluation used the CAMUS dataset (200 apical frames from 50 patients) and reader-verified MIMIC-IV-ECHO frames spanning all 11 views and spectral Doppler signals. Internal metrics were computed on originally annotated test frames and external metrics on single frames as sampled; boxes propagated across clip frames were used for training only.

**Table 1.** Characteristics of the study dataset by split.

| Variable | **Train** | **Validation** | **Test** | **Overall** |
|---|---|---|---|---|
| Studies (patients), n | 94 | 20 | 22 | 136 |
| Age, years, median [IQR] | 73 [67–78] | 74.5 [70–81] | 74.5 [67–81] | 73 [67–79] |
| Male sex, n (%) | 78 (83) | 19 (95) | 18 (82) | 115 (85) |
| **B-mode** |  |  |  |  |
| Annotated frames, n | 699 | 163 | 173 | 1,035 |
| Annotated objects, n | 4,040 | 899 | 973 | 5,912 |
| **Doppler** |  |  |  |  |
| Patients, n | 91 | 20 | 21 | 132 |
| Annotated frames, n | 1,155 | 331 | 320 | 1,806 |
| Annotated objects, n | 2,699 | 772 | 748 | 4,219 |
| **External validation (MIMIC-IV-ECHO)** |  |  |  |  |
| B-mode frames (studies), n | – | – | 44 (44) | – |
| B-mode detections judged, n | – | – | 220 | – |
| Spectral Doppler frames (studies), n | – | – | 40 (40) | – |
| Doppler detections judged (operating point), n | – | – | 94 | – |
| **External validation (CAMUS)** |  |  |  |  |
| Patients, n | – | – | 50 | – |
| Frames (A2C / A4C; ED and ES), n | – | – | 200 | – |
| Annotated objects, n | – | – | 1,459 | – |

B-mode objects are bounding-box annotations on one representative frame per clip. Dash (–), not applicable. IQR, interquartile range; A2C and A4C, apical two- and four-chamber views; ED, end-diastole; ES, end-systole. Per-view and per-signal-class annotation counts are given in Supplementary Table 1. MIMIC-IV-ECHO frames were sampled at random and have no reference annotation; their detections were judged by physicians (Methods).

### 1. General and medical vision models do not localize cardiac structures

To test whether structure localization emerges from general or medical pretraining, we evaluated three open-weight models on the same frames as EchoLens: Qwen2.5-VL-72B^23^, MedGemma-4B^24^ and Grounding DINO^15^ (Methods). At a matched operating point of one box per structure, these models recalled 0% to 3.1% of reference structures in the internal test set (recall of the top-ranked box at an intersection over union (IoU) of 0.5 or higher), compared with 86.5% (95% CI 84.0 to 89.0) for EchoLens (Fig. 2a; Supplementary Tables 2 to 4); the same ordering held on CAMUS against script-generated references (Supplementary Note 1). Every baseline scored below a per-class prior box that never sees the image (fitted on the validation split: 49.1%; fitted on the test set itself: 53.0%). Their boxes did not track image content: deviations of predicted from prior boxes were nearly uncorrelated with the true deviations (r at most 0.13 internally and 0.11 on CAMUS, versus 0.49 to 0.90 and 0.29 to 0.70 for EchoLens; Fig. 2b). MedGemma-4B returned 60 distinct boxes across 1,006 internal queries, and Qwen2.5-VL-72B declined to localize 19.6% of internal queries, almost all for structures present in the frame. MedGemma-4B also failed to localize a bright rectangle on a black synthetic frame under every coordinate convention tested (Supplementary Table 5), so its result reflects an absence of box localization rather than an echocardiography-specific failure.

**Figure 1.**
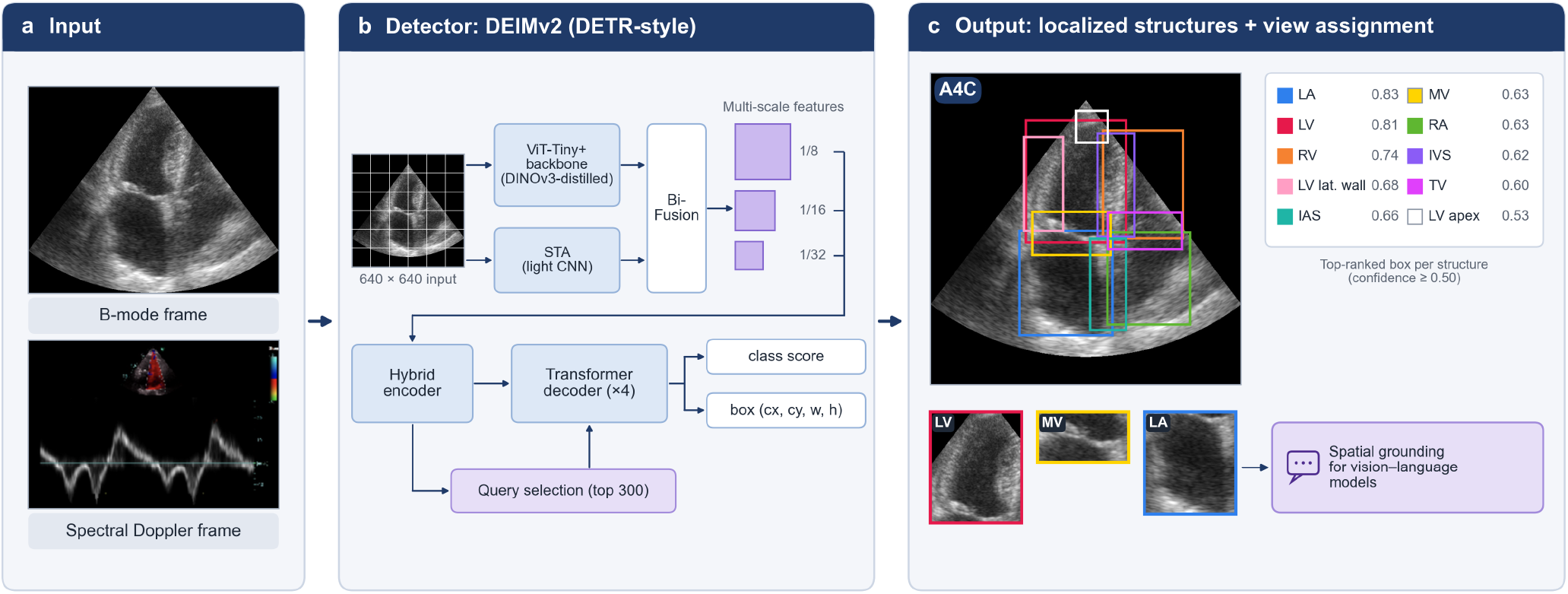
Overview of the EchoLens pipeline. (a) Input: a single transthoracic echocardiography frame, either B-mode (11 views) or spectral Doppler (11 signal types). (b) Detector: DEIMv2, a DETR-style detection transformer. A 640 × 640 preprocessed frame is processed by a ViT-Tiny+ backbone distilled from DINOv3, in parallel with a lightweight convolutional spatial tuning adapter (STA) that extracts fine detail; a hybrid encoder refines and fuses these feature maps, the 300 highest-scoring encoder positions are selected as object queries, and a transformer decoder predicts a class score and a box (centre x, centre y, width, height) for each query. (c) Output: the highest-confidence box of each structure votes for its view, weighted by its confidence, and the view with the highest summed confidence is assigned to the frame. The example is an apical four-chamber frame from the external CAMUS dataset; boxes show the top-ranked prediction of each structure with a confidence of 0.50 or higher (confidences in the key), with crops of the left ventricle, mitral valve and left atrium below. The localized structures provide spatial grounding for vision–language models, the intended application of EchoLens. LA, left atrium; LV, left ventricle; RV, right ventricle; RA, right atrium; IAS, interatrial septum; IVS, interventricular septum; MV, mitral valve; TV, tricuspid valve; A4C, apical four-chamber view.

**Figure 2.**
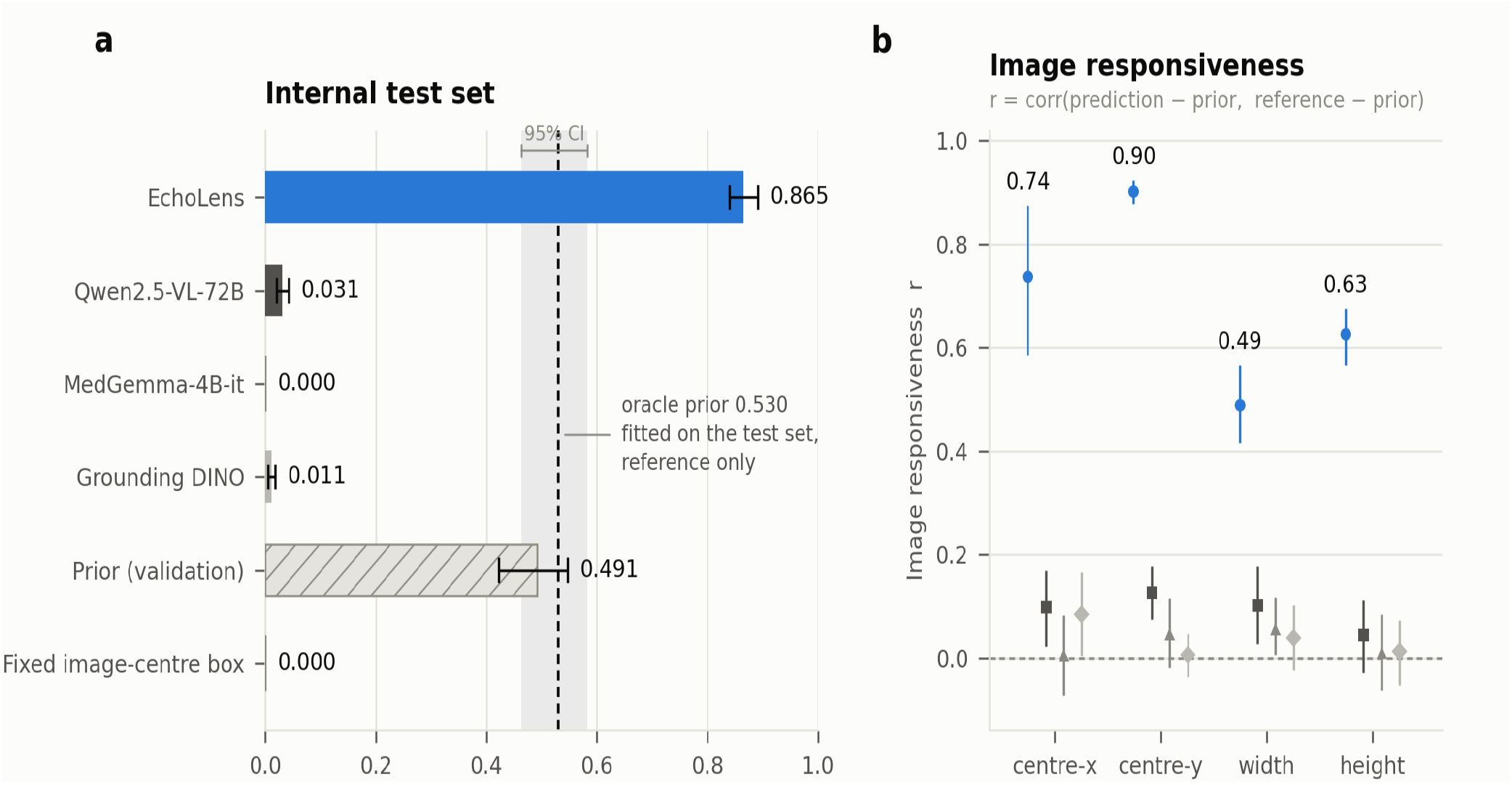
Structure localization by EchoLens and by general-purpose and medical vision models. (a) Recall at IoU ≥ 0.5 of the top-1 box per structure on the internal test set; error bars, 95% CIs from a cluster bootstrap over the 22 test studies. Dashed line and shaded band, oracle prior box per structure-view class derived from the evaluated set itself (reference only). Class prior, median ground-truth box per structure-view class in the validation split; image-centre box, a fixed central box. (b) Image responsiveness: correlation between the deviation of each predicted box from the class prior and the deviation of the reference box from the prior, for box centre (x and y), width and height; values near zero indicate boxes that do not track image content.

### 2. Structure detection across views

Detection of the same structure varied with the view (Fig. 3; all 54 structure-view classes in Supplementary Table 6 and precision–recall curves in Supplementary Fig. 1). Chambers were detected consistently, the left ventricle with an average precision at an IoU of 0.5 (AP50) of 0.86 to 1.00 across seven views and the left atrium with 0.83 to 1.00 across six, whereas smaller structures depended more on the view, most clearly the tricuspid valve (A4C 0.72, Subcostal-4C 0.26, PSAX-AV 0.14), the right ventricular free wall (PSAX-AV 0.92, A4C 0.48) and the right atrium (A4C 0.95, Subcostal-4C 0.57).

**Figure 3.**
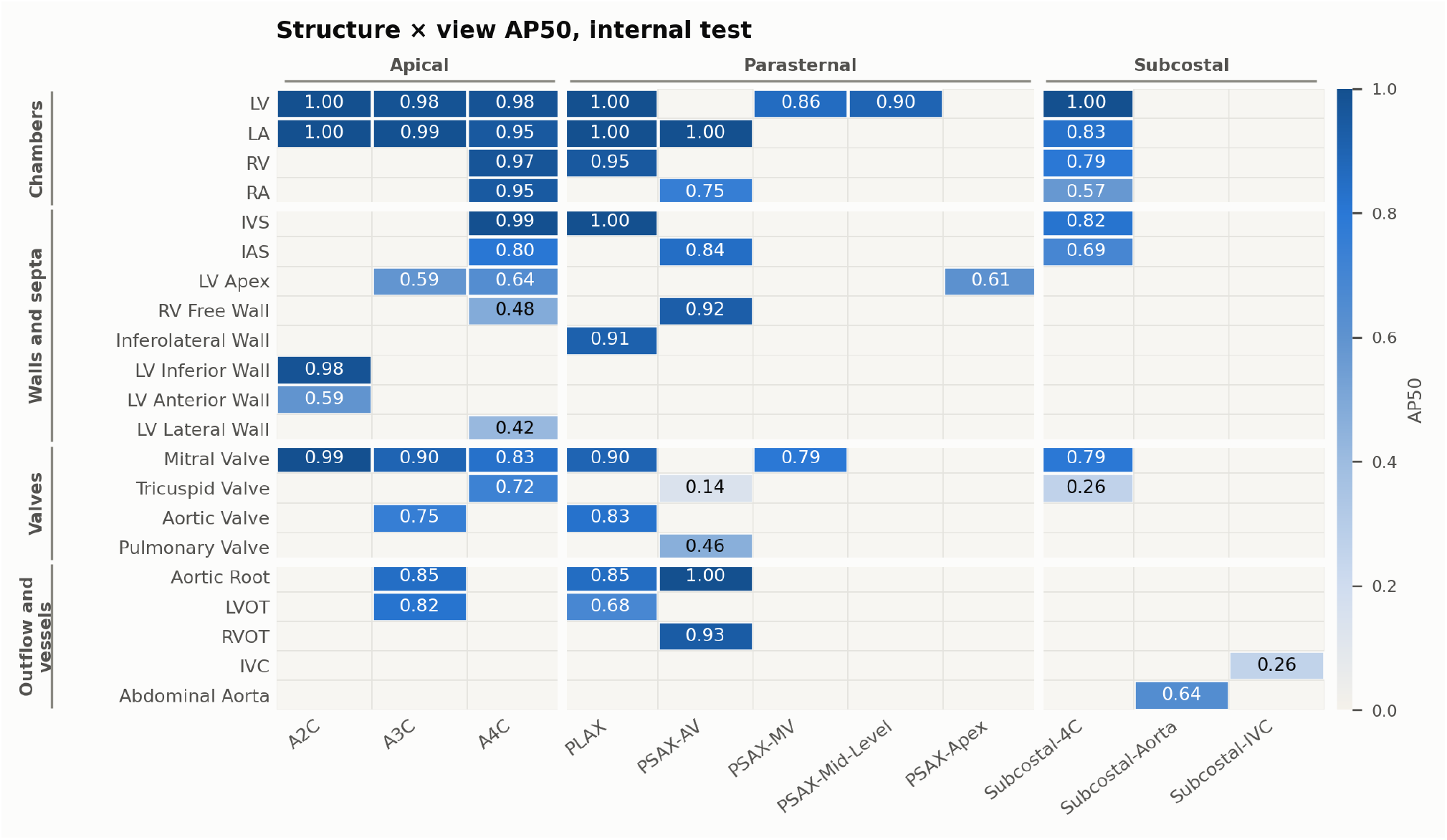
Localization of the same structure in different views (AP50, internal test set, all raw detections). Empty cells: structure not annotated in the view. View abbreviations are defined in Methods.

Pooled across views, EchoLens detected the 21 structures with a mean AP50 of 0.75 (Table 2). AP fell steeply at stricter IoU thresholds (AP75, at an IoU of 0.75, 0.18; AP50:95, averaged over IoU 0.50 to 0.95, 0.30) and was lowest for the inferior vena cava (0.27), the tricuspid valve (0.38) and the left ventricular lateral wall (0.39).

**Table 2.**
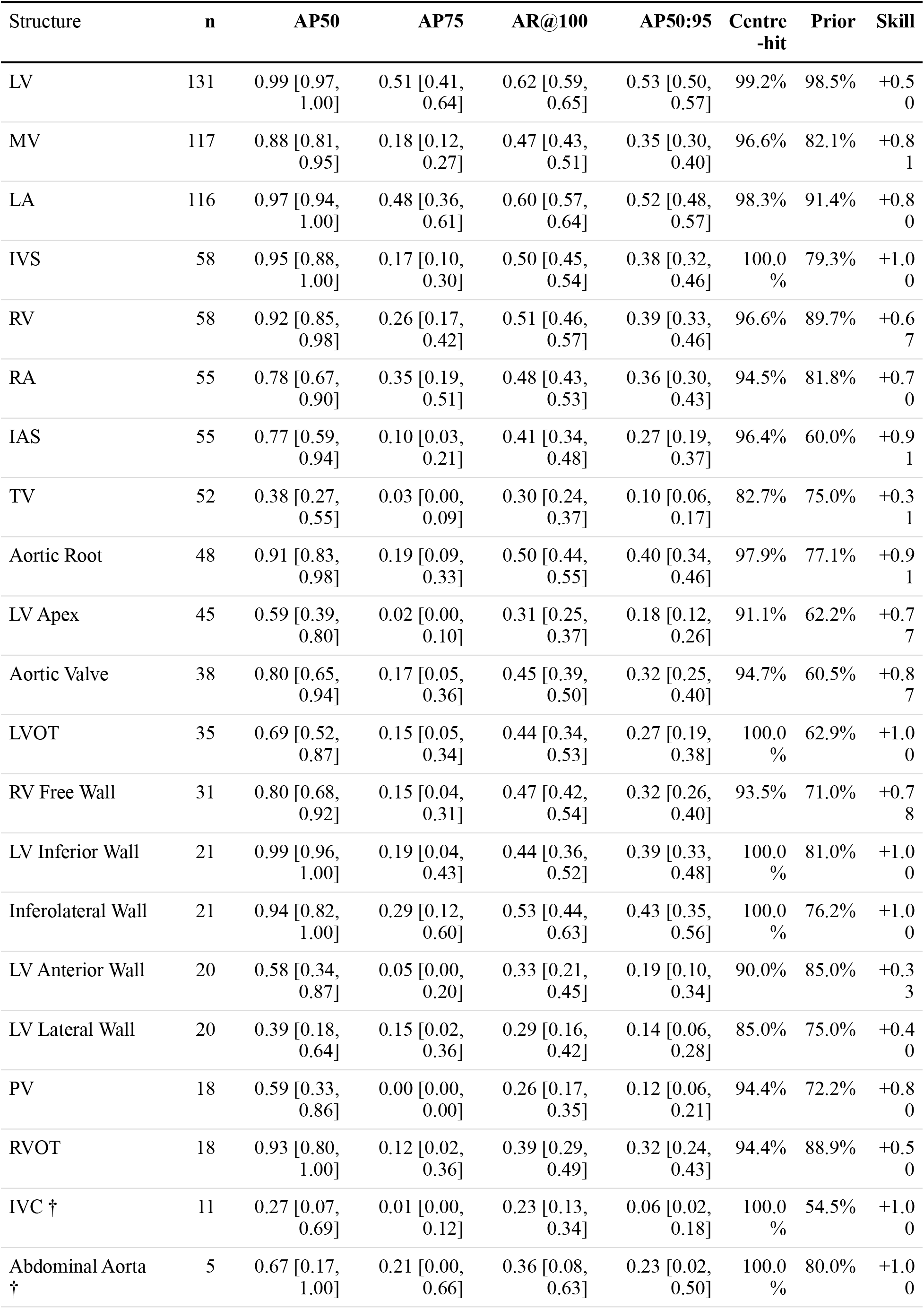
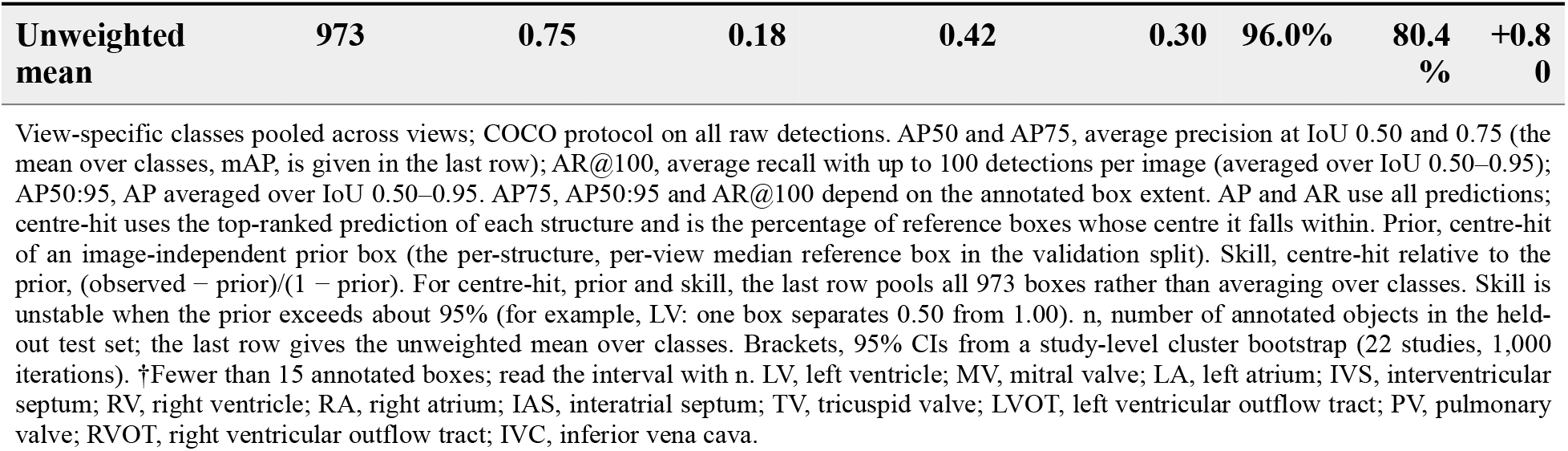
Structure-level detection on the internal test set.

| Structure | **n** | **AP50** | **AP75** | **AR@100** | **AP50:95** | **Centre-hit** | **Prior** | **Skill** |
|---|---|---|---|---|---|---|---|---|
| LV | 131 | 0.99 [0.97, 1.00] | 0.51 [0.41, 0.64] | 0.62 [0.59, 0.65] | 0.53 [0.50, 0.57] | 99.2% | 98.5% | +0.50 |
| MV | 117 | 0.88 [0.81, 0.95] | 0.18 [0.12, 0.27] | 0.47 [0.43, 0.51] | 0.35 [0.30, 0.40] | 96.6% | 82.1% | +0.81 |
| LA | 116 | 0.97 [0.94, 1.00] | 0.48 [0.36, 0.61] | 0.60 [0.57, 0.64] | 0.52 [0.48, 0.57] | 98.3% | 91.4% | +0.80 |
| IVS | 58 | 0.95 [0.88, 1.00] | 0.17 [0.10, 0.30] | 0.50 [0.45, 0.54] | 0.38 [0.32, 0.46] | 100.0% | 79.3% | +1.00 |
| RV | 58 | 0.92 [0.85, 0.98] | 0.26 [0.17, 0.42] | 0.51 [0.46, 0.57] | 0.39 [0.33, 0.46] | 96.6% | 89.7% | +0.67 |
| RA | 55 | 0.78 [0.67, 0.90] | 0.35 [0.19, 0.51] | 0.48 [0.43, 0.53] | 0.36 [0.30, 0.43] | 94.5% | 81.8% | +0.70 |
| IAS | 55 | 0.77 [0.59, 0.94] | 0.10 [0.03, 0.21] | 0.41 [0.34, 0.48] | 0.27 [0.19, 0.37] | 96.4% | 60.0% | +0.91 |
| TV | 52 | 0.38 [0.27, 0.55] | 0.03 [0.00, 0.09] | 0.30 [0.24, 0.37] | 0.10 [0.06, 0.17] | 82.7% | 75.0% | +0.31 |
| Aortic Root | 48 | 0.91 [0.83, 0.98] | 0.19 [0.09, 0.33] | 0.50 [0.44, 0.55] | 0.40 [0.34, 0.46] | 97.9% | 77.1% | +0.91 |
| LV Apex | 45 | 0.59 [0.39, 0.80] | 0.02 [0.00, 0.10] | 0.31 [0.25, 0.37] | 0.18 [0.12, 0.26] | 91.1% | 62.2% | +0.77 |
| Aortic Valve | 38 | 0.80 [0.65, 0.94] | 0.17 [0.05, 0.36] | 0.45 [0.39, 0.50] | 0.32 [0.25, 0.40] | 94.7% | 60.5% | +0.87 |
| LVOT | 35 | 0.69 [0.52, 0.87] | 0.15 [0.05, 0.34] | 0.44 [0.34, 0.53] | 0.27 [0.19, 0.38] | 100.0% | 62.9% | +1.00 |
| RV Free Wall | 31 | 0.80 [0.68, 0.92] | 0.15 [0.04, 0.31] | 0.47 [0.42, 0.54] | 0.32 [0.26, 0.40] | 93.5% | 71.0% | +0.78 |
| LV Inferior Wall | 21 | 0.99 [0.96, 1.00] | 0.19 [0.04, 0.43] | 0.44 [0.36, 0.52] | 0.39 [0.33, 0.48] | 100.0% | 81.0% | +1.00 |
| Inferolateral Wall | 21 | 0.94 [0.82, 1.00] | 0.29 [0.12, 0.60] | 0.53 [0.44, 0.63] | 0.43 [0.35, 0.56] | 100.0% | 76.2% | +1.00 |
| LV Anterior Wall | 20 | 0.58 [0.34, 0.87] | 0.05 [0.00, 0.20] | 0.33 [0.21, 0.45] | 0.19 [0.10, 0.34] | 90.0% | 85.0% | +0.33 |
| LV Lateral Wall | 20 | 0.39 [0.18, 0.64] | 0.15 [0.02, 0.36] | 0.29 [0.16, 0.42] | 0.14 [0.06, 0.28] | 85.0% | 75.0% | +0.40 |
| PV | 18 | 0.59 [0.33, 0.86] | 0.00 [0.00, 0.00] | 0.26 [0.17, 0.35] | 0.12 [0.06, 0.21] | 94.4% | 72.2% | +0.80 |
| RVOT | 18 | 0.93 [0.80, 1.00] | 0.12 [0.02, 0.36] | 0.39 [0.29, 0.49] | 0.32 [0.24, 0.43] | 94.4% | 88.9% | +0.50 |
| IVC † | 11 | 0.27 [0.07, 0.69] | 0.01 [0.00, 0.12] | 0.23 [0.13, 0.34] | 0.06 [0.02, 0.18] | 100.0% | 54.5% | +1.00 |
| Abdominal Aorta † | 5 | 0.67 [0.17, 1.00] | 0.21 [0.00, 0.66] | 0.36 [0.08, 0.63] | 0.23 [0.02, 0.50] | 100.0% | 80.0% | +1.00 |
| **Unweighted mean** | **973** | **0.75** | **0.18** | **0.42** | **0.30** | **96.0%** | **80.4%** | **+0.80** |

View-specific classes pooled across views; COCO protocol on all raw detections. AP50 and AP75, average precision at IoU 0.50 and 0.75 (the mean over classes, mAP, is given in the last row); AR@100, average recall with up to 100 detections per image (averaged over IoU 0.50–0.95); AP50:95, AP averaged over IoU 0.50–0.95. AP75, AP50:95 and AR@100 depend on the annotated box extent. AP and AR use all predictions; centre-hit uses the top-ranked prediction of each structure and is the percentage of reference boxes whose centre it falls within. Prior, centre-hit of an image-independent prior box (the per-structure, per-view median reference box in the validation split). Skill, centre-hit relative to the prior, (observed − prior)/(1 − prior). For centre-hit, prior and skill, the last row pools all 973 boxes rather than averaging over classes. Skill is unstable when the prior exceeds about 95% (for example, LV: one box separates 0.50 from 1.00). n, number of annotated objects in the held-out test set; the last row gives the unweighted mean over classes. Brackets, 95% CIs from a study-level cluster bootstrap (22 studies, 1,000 iterations). †Fewer than 15 annotated boxes; read the interval with n. LV, left ventricle; MV, mitral valve; LA, left atrium; IVS, interventricular septum; RV, right ventricle; RA, right atrium; IAS, interatrial septum; TV, tricuspid valve; LVOT, left ventricular outflow tract; PV, pulmonary valve; RVOT, right ventricular outflow tract; IVC, inferior vena cava.

Low AP did not reflect failure to find these structures: the centre of EchoLens’s top-ranked box fell within the reference box in 96.0% of cases, against 80.4% for an image-independent prior, and walls were found as often as other structures (95.9% vs 96.0%; difference −0.1 percentage points, 95% CI −2.9 to +2.6; Table 2 and Supplementary Table 7; examples in Supplementary Fig. 2). The reproducibility analysis below attributes the loss at strict thresholds to the box annotation protocol.

Because predictions were pooled by structure irrespective of their view label, structure detection did not depend on a view decision, and 1.4% of correct detections (12 of 842) carried the label of another view. Of detections naming a structure not annotated in the frame, 62% named a structure of a neighbouring view of the same family (Supplementary Table 8). The view implied by the detections matched the reference in 94% of frames (162 of 173, 11 views; Supplementary Fig. 3).

At a confidence threshold of 0.50, selected by maximizing F1 on the validation set, precision was 84.1% (95% CI 81.2 to 86.9) and recall 84.9% (82.3 to 87.5) on the test set, close to validation (83.1% and 83.0%), and F1 stayed within 1 percentage point of its maximum from 0.38 to 0.62 (Supplementary Fig. 4).

### 3. Reproducibility of box extent

To establish whether low strict-overlap performance reflected the detector or the annotation target, a second experienced reader independently re-annotated 140 test frames from nine views (Methods). The reader placed boxes at the same locations as the reference but drew them smaller (a median of 0.50 of the reference area), so only 37% of the reader’s boxes reached an IoU of 0.5 with the reference, fewer than for an image-independent prior (47%) and EchoLens (87%; Table 3). After a single global scale correction fitted on chamber cavities, the reader reproduced EchoLens’s ordering across structure classes, with larger deficits than EchoLens for walls and the apex. Box centres fell within the reference box equally often for the reader and EchoLens (95.3% and 95.7%, against 79.7% for the prior), and the reader did not identify 35 of 921 annotated structures (3.8%), including 10 of 18 pulmonary valves.

**Table 3.** Second-reader agreement with the reference annotation.

|  | EchoLens | Anatomical prior | Second reader, as drawn | Second reader, scale-corrected | EchoLens skill | Reader skill |
|---|---|---|---|---|---|---|
| **Mean IoU, by structure class** |  |  |  |  |  |  |
| Chamber cavity (n = 322) | 0.744 | 0.558 | 0.478 | 0.644 |  |  |
| Wall / septum (n = 217) | 0.665 | 0.425 | 0.425 | 0.540 |  |  |
| Valve (n = 187) | 0.623 | 0.409 | 0.387 | 0.525 |  |  |
| Vessel / outflow (n = 117) | 0.664 | 0.364 | 0.470 | 0.512 |  |  |
| Apical region (n = 43) | 0.562 | 0.358 | 0.228 | 0.291 |  |  |
| **Boxes reaching an IoU threshold** |  |  |  |  |  |  |
| IoU ≥ 0.3 | 95% | 75% | 78% | 89% |  |  |
| IoU ≥ 0.4* | 93% | 61% | 58% | 81% |  |  |
| IoU ≥ 0.5* | 87% | 47% | 37% | 69% |  |  |
| IoU ≥ 0.7* | 53% | 14% | 7% | 25% |  |  |
| IoU ≥ 0.75* | 40% | 8% | 3% | 15% |  |  |
| **Centre-hit, by structure class†** |  |  |  |  |  |  |
| Chamber cavity | 97.5% | 92.5% | 96.3% | 96.3% | 0.667 | 0.500 |
| Wall / septum | 95.9% | 74.2% | 93.5% | 93.5% | 0.839 | 0.750 |
| Valve | 92.0% | 73.3% | 93.0% | 93.0% | 0.700 | 0.740 |
| Vessel / outflow tract | 98.3% | 71.8% | 98.3% | 98.3% | 0.939 | 0.939 |
| Apical region | 90.7% | 60.5% | 97.7% | 97.7% | 0.765 | 0.941 |
| **All structures** | **95.7%** | **79.7%** | **95.3%** | **95.3%** | **0.789** | **0.767** |

140 internal test frames; 886 boxes matched to a reference box. Mean IoU and centre-hit against the reference annotation. Second reader, scale-corrected, applies a single global width and height factor (×1.442, ×1.329) fitted on chamber-cavity boxes only, so every other class is out of sample. Anatomical prior, per-structure, per-view median reference box fitted on the validation split, which never sees the image. Skill = (observed − prior) / (1 − prior); 1.00 is perfect given the prior and 0.00 is no better than not looking at the image. \*Rows in which the second reader scored below the image-independent prior. †Centre-hit asks whether the box centre falls inside the reference box; it is scale-invariant, so the two reader columns are identical by construction.

**Table 4.** Doppler signal detection on the internal test set.

| Doppler signal class | n | **AP50** | **AP75** | **AR@100** | **AP50:95** |
|---|---|---|---|---|---|
| TDI, tricuspid valve lateral annulus | 52 | 0.77 [0.62, 0.90] | 0.60 [0.39, 0.78] | 0.69 [0.64, 0.73] | 0.48 [0.36, 0.59] |
| TDI, mitral valve | 99 | 0.78 [0.64, 0.90] | 0.29 [0.19, 0.43] | 0.59 [0.55, 0.63] | 0.36 [0.29, 0.45] |
| PW, mitral inflow | 88 | 0.98 [0.96, 1.00] | 0.74 [0.60, 0.90] | 0.74 [0.71, 0.78] | 0.63 [0.58, 0.70] |
| PW, LVOT | 115 | 0.97 [0.94, 1.00] | 0.51 [0.38, 0.69] | 0.69 [0.65, 0.73] | 0.54 [0.48, 0.62] |
| PW, abdominal aortic flow | 19 | 0.98 [0.95, 1.00] | 0.52 [0.26, 0.83] | 0.64 [0.58, 0.71] | 0.54 [0.45, 0.63] |
| CW, tricuspid regurgitant flow | 146 | 0.89 [0.83, 0.95] | 0.70 [0.59, 0.81] | 0.74 [0.70, 0.78] | 0.60 [0.53, 0.67] |
| CW, pulmonary regurgitant flow † | 18 | 0.65 [0.35, 0.93] | 0.25 [0.05, 0.62] | 0.67 [0.56, 0.79] | 0.38 [0.19, 0.60] |
| CW, pulmonary forward flow | 31 | 0.77 [0.59, 0.94] | 0.54 [0.30, 0.79] | 0.70 [0.65, 0.75] | 0.51 [0.38, 0.64] |
| CW, mitral regurgitant flow | 29 | 0.63 [0.35, 0.85] | 0.40 [0.14, 0.71] | 0.64 [0.50, 0.81] | 0.36 [0.20, 0.55] |
| CW, aortic regurgitant flow | 33 | 0.93 [0.81, 1.00] | 0.63 [0.35, 0.93] | 0.64 [0.56, 0.73] | 0.52 [0.38, 0.67] |
| CW, aortic outflow | 118 | 0.93 [0.87, 0.97] | 0.72 [0.60, 0.85] | 0.70 [0.65, 0.76] | 0.59 [0.53, 0.66] |
| Unweighted mean | 748 | 0.84 | 0.54 | 0.68 | 0.50 |

Medial and lateral mitral TDI merged into a single class; COCO protocol on all raw detections. TDI, tissue Doppler imaging; PW, pulsed-wave; CW, continuous-wave; LVOT, left ventricular outflow tract. Metrics as in Table 2; n, number of annotated objects in the held-out test set; the last row gives the unweighted mean over classes. Brackets, 95% CIs from a frame-level bootstrap (1,000 iterations), as the Doppler data carry no study identifier. †Fewer than 5 contributing frames at some IoU thresholds.

### 4. Doppler signal detection

The Doppler detector identified the 11 signal types with a mean AP50 of 0.84 (Table 4). AP75 was far higher than for B-mode structures (0.54 vs 0.18), consistent with a spectral envelope being a more reproducible box than an anatomical structure, and the regurgitant signals of the mitral (AP50 0.63) and pulmonic (0.65) valves were the weakest classes.

The top-ranked detection identified the signal type in 90.6% (95% CI 87 to 93) of test frames, and its box reached an IoU of 0.5 or higher with an annotated beat for 94.8% (92 to 97) of signal classes, against 9.2% for a constant box per class. At the validation-selected operating point (the top-ranked signal class, then every detection of that class scoring 0.36 or higher), precision was 85.8% (83 to 88) and recall 83.0%. Mitral regurgitation was the most confused class: 7 of 13 frames were assigned to tricuspid regurgitation, whereas no tricuspid regurgitation frame (0 of 63) was assigned to mitral regurgitation.

### 5. External validation in MIMIC-IV-ECHO

We applied EchoLens to 44 MIMIC-IV-ECHO frames, one per study and four from each of the 11 B-mode views, with the internal protocol of up to five structures per frame, and two experts independently judged each reported structure (Methods). The first and second physicians judged 84.8% (95% CI 76.3 to 92.1) and 85.3% (77.3 to 92.6) of reported structures correct, respectively, 81.0% when both had to agree and 91.5% when either sufficed (Table 5; Supplementary Table 9), and both judged at least one structure correct in 38 of 44 frames. The physicians agreed on 89.5% of detections (κ 0.56; prevalence-adjusted κ 0.79), with disagreement concentrated on the inferior vena cava (42.9% of its detections, against 7.1% or less for every other structure). Of 40 randomly sampled MIMIC-IV-ECHO spectral frames reviewed by a physician (Methods), six (15.0%; 95% CI 5.0 to 27.5) showed signals outside the model’s ontology and four lacked an adjudicable signal type. In the remaining 30, EchoLens identified the signal type in 83.3% (69.0 to 96.6), against 90.6% internally, with a precision of 84.0% (70.3 to 95.9) at the validation-selected operating point (Table 5; Supplementary Table 10).

**Table 5.**
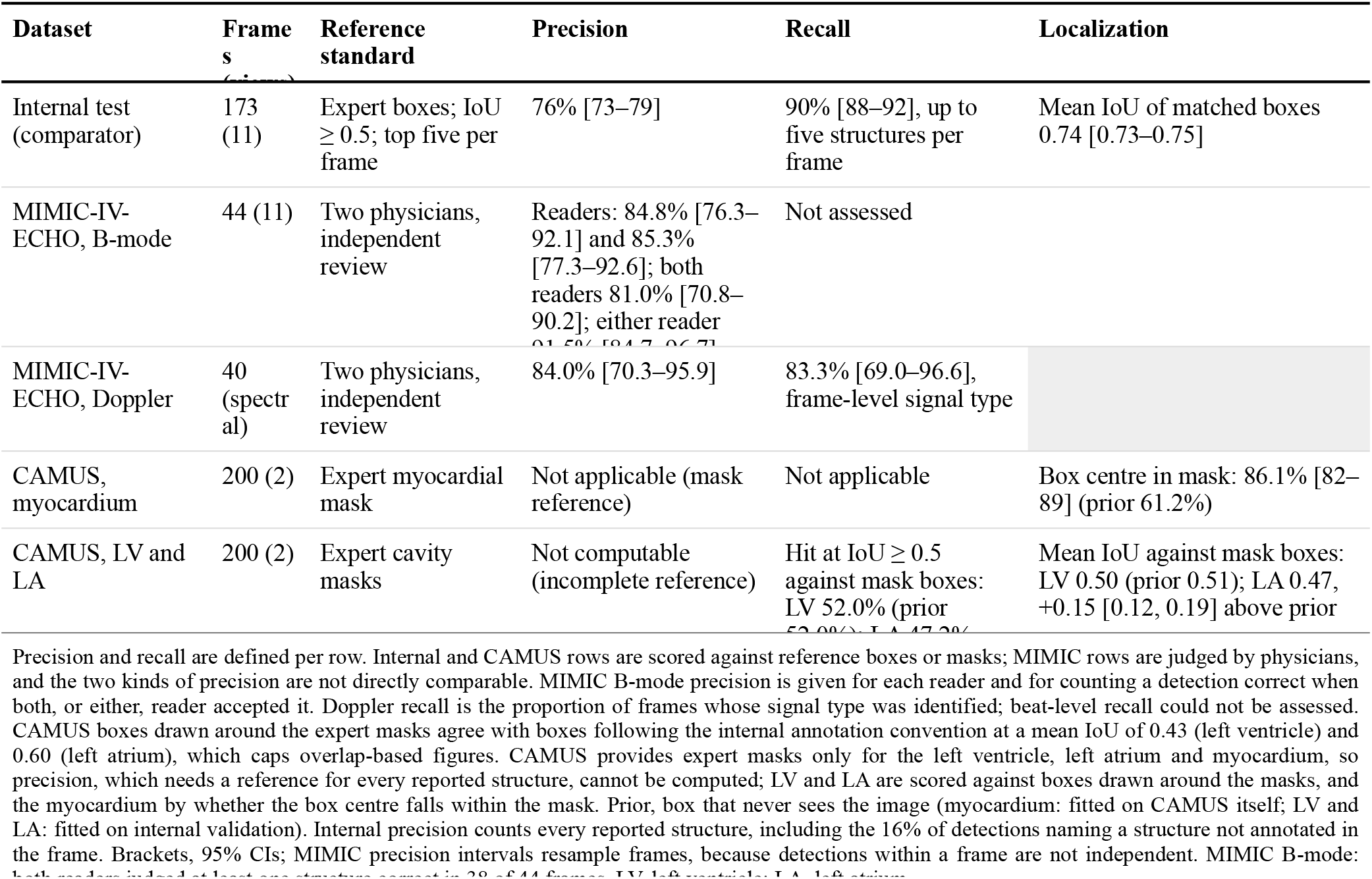
External validation.

### 6. External validation on the CAMUS dataset

CAMUS provides expert masks only for the left ventricular cavity, left atrium and myocardium (Methods). Boxes around these masks follow a different convention from ours: a box drawn to the internal convention was a median of 2.3-fold larger than the box around the left ventricular mask (1.6-fold for the left atrium), and the two references agreed at a mean IoU of only 0.43 and 0.60 (Supplementary Table 11). Against the mask boxes, EchoLens exceeded an image-independent prior for the left atrium (mean IoU difference +0.15 [0.12, 0.19]) but not for the left ventricle, whose position is nearly fixed in the curated apical views of CAMUS (−0.02 [−0.05, +0.02]; Supplementary Table 12). The myocardium gave the clearest external test: the centres of EchoLens’s wall boxes fell within the expert myocardial mask in 86.1% (95% CI 82 to 89) of cases, against 61.2% for a prior fitted on CAMUS itself and 34.3% for one fitted on internal data (Table 5; Supplementary Table 13). Structures without a native reference are reported against script-generated references in Supplementary Note 1.

## 4. Discussion

EchoLens localized 21 cardiac structures across 11 standard views and identified 11 spectral Doppler signal types after training on 1,035 annotated frames from 136 patients. It localized 86.5% of structures in held-out internal frames and had 81% to 92% of its detections judged correct by two physicians in an external set spanning all 11 views, whereas general-purpose and medical vision-language models and an open-vocabulary detector recalled 3% or fewer. On the internal test set, EchoLens placed its boxes on the correct structure as reliably as a blinded expert reader. To our knowledge, it is the first detection framework to span the standard adult transthoracic examination.

The comparison models did not fail by being imprecise: their boxes did not follow the image. Their deviations from a fixed anatomical prior were uncorrelated with the true deviations (r at most 0.13, versus 0.49 to 0.90 for EchoLens on internal frames), every model scored below that prior, and MedGemma returned a near-constant box per structure. Fine-grained localization therefore did not emerge from scale, medical pretraining or open-vocabulary detection training, even when the view and an anatomical definition were supplied; it had to be learned from spatial supervision. The analysis also carries a methodological lesson. Because cardiac anatomy is stereotyped within standard views, a box that never sees the image localized about half of all structures (49% to 53%) at an IoU of 0.5, so claims of visual grounding in echocardiography are interpretable only against image-independent controls. The left ventricle illustrates the problem most clearly. Because the prior is fitted per structure-view class, it is equivalent to a view classifier followed by a lookup of the typical box for that view. Given the view, this lookup placed its centre within the reference left ventricle in 98.5% of internal frames and within the left atrium in 91.4%, compared with 99.2% and 98.3% for EchoLens (Table 2), and on CAMUS, scored against expert masks, it matched EchoLens for the left ventricle. View classifiers already exceed 90% accuracy^2,5,7^, so a detector trained only on large chambers such as the left ventricle and left atrium adds little beyond view classification. Previous left ventricular detection studies have, to our knowledge, not reported such a comparison, and high performance on these structures alone does not show that a model uses the image. The evidence for image-based localization lies in the structures a fixed box cannot find, such as the interatrial septum, valves and walls, where EchoLens recovered most of the gap left by the prior (skill 0.82 for walls and 0.91 for the interatrial septum). Saliency maps such as Grad-CAM, where they can be computed, approximate where a model attended rather than naming and placing a structure, and they localize less accurately than dedicated localization networks^10,11^; explicit detection provides that evidence directly, and in a stereotyped anatomy any regional explanation, saliency included, needs the same image-independent control to be interpretable.

Where EchoLens fell short, the data implicate the reference standard and the images more than the detector (examples in Supplementary Fig. 2). Strict-overlap performance was low (AP75 0.18), but a blinded second reader agreed with the reference less closely than EchoLens did in every structure class, while the box centre fell within the reference box equally often for both (95.3% and 95.7%). Box extent is thus a convention that experts do not reproduce, and AP75 operates below the reproducibility of the protocol that defines it; because EchoLens learned the reference annotators’ convention, this shows that box extent is convention-dependent, not that EchoLens exceeds human performance. The remaining weaknesses followed anatomy and acquisition. Walls, despite the lowest AP of any structure class, were found as reliably as other structures (box centre within the reference in 95.9% vs 96.0%), so their deficit lies in box extent rather than in finding the wall; the tricuspid valve, not a wall, had the lowest skill relative to an image-independent prior. The pulmonary valve could not be identified even by the second reader in 10 of 18 frames, and the inferior vena cava, although well localized by its top-ranked box (vessel centre-hit 98.3%), generated false-positive and vessel-confused detections in subcostal frames. Of the internal detections that matched no annotated structure, most (62%) named a structure belonging to a neighbouring view, so errors arose mainly in the view or level assignment rather than in identifying the structure itself. Because a well-placed box carrying the wrong structure name is more likely to be believed than a misplaced one, such errors should be filtered at the view or level assignment, where they arise, before detections are used downstream.

Performance transferred to external data, with the reference standard shaping what could be measured. Two echocardiographers judged 81% to 92% of EchoLens’s detections across the 11 views of MIMIC-IV-ECHO correct, and on CAMUS the centres of wall boxes fell within the expert myocardial mask in 86% of cases, against 61% for a prior fitted on CAMUS itself and 34% for one fitted on internal data, indicating localization from image content rather than typical position. CAMUS also shows how much the reference matters: boxes around its expert masks agreed with boxes following our annotation convention at a mean IoU of only 0.43 to 0.60, and for the left ventricle, whose position is nearly fixed in the curated apical views of CAMUS, a constant box matched EchoLens.

Doppler behaved differently from B-mode. Spectral envelopes are small and move along the time axis, so a constant box localized 9% of signals against 95% for EchoLens, and signal-type identification transferred to external data (83.3% vs 90.6%). The dominant error was one-directional: mitral regurgitation was assigned to tricuspid regurgitation in 7 of 13 frames and never the reverse. These two systolic, high-velocity envelopes are distinguished mainly by timing and clinical context, and mitral regurgitation was the less frequent class in the training data.

Previous detection work in echocardiography has localized one or a few targets, typically the left ventricle or the chambers in a single view^16–20,22^ (Supplementary Table 14). EchoLens extends detection to the standard examination. Its outputs, labelled boxes with confidence scores, are the format vision-language models already use for grounding. The same detections record which structures are visible in each frame, which could support acquisition quality control and tie automated measurements to the structures they describe. We did not attempt segmentation: pixel-level delineation of every structure is not part of routine echocardiographic reporting and would be far costlier to annotate for thin and small structures such as valves and walls. Because vision-language models learn to ground from box-level supervision, which expert annotation cannot provide at the scale of their training data, EchoLens can also generate grounding labels across large unannotated archives; whether such labels improve the accuracy and verifiability of generated reports is the next question to test.

This study has limitations. Development data came from one center and 136 patients, 85% of them men, with a median age of 73 years and none younger than 48. External evaluation of multiple views and of Doppler relied on modest reader-adjudicated samples, and CAMUS provides native references for only three structures. Reproducibility was assessed with a single second reader; agreement among multiple experts would better define the ceiling of the annotation protocol. Training labels propagated across clips are approximate, although all evaluation used originally annotated frames. The ontology excludes colour Doppler, M-mode, several views and pathological findings, axis-aligned boxes represent curved and thin structures imperfectly, and EchoLens operates on single frames. The comparison models were open-weight so that protected data could be evaluated identically, and the medical model was the 4-billion-parameter MedGemma. We did not fine-tune vision-language models on our annotations; the comparison shows that structure localization does not arise from pretraining, not that EchoLens would outperform a vision-language model trained for the task.

Fine-grained structure localization did not emerge from current general or medical vision-language models, but it was learned from a modest set of expert-annotated frames and transferred across views and datasets. By making explicit where each structure lies, EchoLens provides the spatial layer that visually grounded echocardiographic interpretation requires.

## 5. Methods

### Study population

We included 136 transthoracic echocardiographic studies, each from a different patient, randomly selected from consecutive studies acquired at Mayo Clinic between November 2017 and December 2019. Studies were assigned at the patient level to training, validation and test sets (70:15:15; Table 1).

### Annotation

A single representative frame per clip was annotated by eight annotators with medical degrees, trained in echocardiographic anatomy, on the MD.ai platform (MD.ai Inc., New York, NY, USA) following a predefined protocol (Supplementary Note 2). B-mode annotations comprised 5,912 bounding boxes of 21 cardiac structures across 11 standard views (parasternal long-axis, PLAX; parasternal short-axis at the aortic valve, mitral valve, mid-ventricular and apical levels, PSAX-AV, PSAX-MV, PSAX-Mid-Level and PSAX-Apex; apical two-, three- and four-chamber, A2C, A3C and A4C; and subcostal four-chamber, inferior vena cava and abdominal aorta, Subcostal-4C, Subcostal-IVC and Subcostal-Aorta); each structure-view pair (for example, the left ventricle [LV] in A4C) was a separate class, giving 54 classes. Spectral Doppler annotations comprised 4,219 boxes on 1,806 frames, each enclosing one beat of a spectral envelope. Medial and lateral mitral annular TDI were merged because they cannot be distinguished from the spectral display alone, giving 11 Doppler classes. Per-view and per-class counts are given in Supplementary Table 1.

### Preprocessing

After annotation, Digital Imaging and Communications in Medicine (DICOM) pixel data were converted to RGB and the ultrasound sector was isolated by masking regions outside it, including burned-in patient identifiers; for B-mode, the sector was defined as the pixels that were non-zero in at least one frame of the clip, on-screen text was removed by eroding this mask in the upper three quarters of the image and by masking static writing in the upper-left corner, and the electrocardiogram (ECG) trace was retained. Spectral Doppler frames were processed by removing the top information bar and corner annotations, masking regions outside the spectral display, and cropping and rescaling the spectral region; the ECG trace was retained and on-screen text removed. Frames were resized to 640 × 640 pixels after zero-padding to a square (letterboxing, without cropping) with bicubic interpolation and normalized with ImageNet channel statistics (mean 0.485, 0.456, 0.406; standard deviation 0.229, 0.224, 0.225). External frames received the same preprocessing, with no adaptation or fine-tuning.

### Model development

Boxes annotated on one frame per clip were propagated to the remaining frames of the clip for training only, expanding the B-mode training data to 364,894 object instances; Doppler frames were not propagated. Separate B-mode and Doppler detectors were trained with DEIMv2^25^, a real-time detection transformer with one-to-one matching and a matchability-aware loss, in its M configuration with a ViT-Tiny+ backbone distilled from the self-supervised DINOv3 model^26^. Both detectors used an input size of 640 × 640, AdamW (learning rate 5 × 10⁻⁴, and 2.5 × 10⁻⁵ for the backbone; weight decay 10⁻⁴; gradient clipping at 0.1), a total batch size of 32 across four GPUs with synchronized batch normalization, mixed precision and an exponential moving average of the weights, and up to 102 epochs with mosaic, photometric distortion, zoom-out, IoU-based cropping and horizontal flip augmentation. Training took about 23 h for B-mode (15.6 min per epoch) and 30 min for Doppler on four NVIDIA A100 GPUs; full settings are in Supplementary Note 3. The checkpoint with the highest validation AP during the first training stage was used. All evaluation used originally annotated frames only. Inference took 20.7 ms per frame for B-mode and 21.0 ms for Doppler on one NVIDIA A100 GPU (batch size 1, model forward pass and post-processing).

### Evaluation

#### Structure-level detection

EchoLens returns the 300 highest-scoring class predictions per image, all of which were evaluated. Each prediction, labelled as a structure-view pair, was mapped to its anatomical structure irrespective of its view label (i.e., LV from each view was pooled together), and non-maximum suppression at IoU 0.7 was applied within each structure. Detection was evaluated with the COCO protocol^27^ over the 21 structures. AP50 was the primary measure, because grounding requires a box that identifies and contains the correct structure, not one that reproduces the annotator’s exact edges. AP75 and AP averaged over IoU 0.50 to 0.95 (AP50:95) measure agreement with the annotated extent; they matter for measurement-grade localization, which EchoLens does not claim, and they penalize small shifts of thin structures such as walls, septa and leaflets. Average recall with up to 100 detections per image (AR@100) quantified sensitivity. Mean IoU between each reference box and the top-ranked prediction of the same structure measured agreement in extent. Centre-hit, whether that box’s centre fell within the reference box, measured localization and was always reported against an image-independent prior (the per-structure, per-view median reference box in the validation split) as skill = (observed − prior)/(1 − prior). Distance-based measures were not used because zoom and depth vary across images and datasets.

#### Operating points

For B-mode, a confidence threshold of 0.50 was selected by maximizing F1 at IoU 0.5 on the validation split and applied once to test data. For Doppler, the top-ranked signal class was taken as the frame’s signal type and all detections of that class scoring 0.36 or higher were retained, with the threshold also selected on validation. Selection uncertainty was estimated by re-selecting each threshold on resampled validation frames (Supplementary Table 15).

#### View assignment

For each image, the highest-confidence prediction of each anatomical structure was retained irrespective of its view label, the confidences of the retained predictions were summed by view, and the view with the highest total was assigned. View assignment was not used for structure-level evaluation (accuracy in Supplementary Fig. 3); structure-level AP50 was compared between frames whose assigned view did and did not match the reference view (Supplementary Table 8).

#### Second-reader agreement

An experienced reader re-annotated all 140 test frames from the nine views that define more than two structures (PSAX-MV and PSAX-Mid-Level excluded), blinded to the reference boxes, to EchoLens’s output and to which structures were annotated. Reader and EchoLens boxes were scored against the reference with the same evaluator. For a scale-corrected comparison, a single global width and height factor (×1.442, ×1.329) was fitted on chamber-cavity boxes only, so other classes were evaluated out of sample.

#### Comparison with vision-language and open-vocabulary models

We compared EchoLens with Qwen2.5-VL-72B-Instruct^23^, MedGemma-4B-it^24^ and Grounding DINO (base)^15^, open-weight models that could be applied to protected data, all given the same 640 × 640 frames. The vision-language models were prompted once per structure with the view, a one-sentence anatomical definition and a request for a bounding box in JSON, with prompts frozen on the validation split and deterministic decoding; Grounding DINO was prompted with the structure names of the view (Supplementary Table 5). Coordinate conventions were checked on synthetic images (Supplementary Table 5), and unparseable outputs and absent boxes counted as misses. Because the vision-language models return one box without a confidence score, all models were compared on the top-ranked box per structure-view class, by recall at IoU 0.5 or higher (primary) and mean IoU. Three image-independent controls were included: the validation-fitted prior box, the same prior fitted on the evaluated set (reference only) and a fixed image-centre box. Image responsiveness was measured as the correlation between predicted and reference deviations from the prior box for each box parameter. Spectral Doppler was not part of the test-set comparison: in a preliminary evaluation on internal validation frames, all three models localized Doppler signals no better than a constant box that never sees the image (Supplementary Table 16).

#### External validation

MIMIC-IV-ECHO^28^ data were processed locally under the PhysioNet credentialed data use agreement. For B-mode, 44 frames from 44 studies, four for each of the 11 views, were drawn at random (seed 0) from a pool of 330 frames from 307 studies sampled across the 11 views. EchoLens outputs (up to five structures per frame) were reviewed independently by two blind echocardiographers, who judged each box correct, incorrect or unsure and recorded visible structures that were not detected. Precision is reported per reader and for counting a box correct when both, or either, reader accepted it; agreement was summarized by Cohen’s κ.

For MIMIC-IV-ECHO Doppler, 40 frames were drawn at random from a 200-frame pool of spectral stills from 195 studies. A physician judged each of the top five detections correct, incorrect or undecidable. Because the review interface allowed the signal label to be revised after the detections were shown, the reference signal type and ontology membership were re-established by consensus of two physicians under a written ontology rule, starting from the first physician’s label recorded before the detections were shown and a second physician’s blinded label, and precision is reported without crediting detections that matched a revised label.

CAMUS^29^ contributed 200 apical two- and four-chamber frames (end-diastole and end-systole) from its 50 test patients. Its expert segmentations cover only the left ventricular cavity, left atrium and myocardium. Left ventricular and left atrial reference boxes were drawn tightly around the masks and compared with predictions by mean IoU and hits at IoU 0.5 or higher, against the validation-fitted prior; agreement between mask-derived boxes and boxes following the internal annotation convention was also measured. Wall predictions were evaluated by whether the box centre fell within the myocardial mask and by the fraction of box area on it. Structures without a native reference were excluded; results against script-generated references are reported in Supplementary Note 1.

### Statistical analysis

Ninety-five per cent confidence intervals were estimated by nonparametric cluster bootstrap (1,000 iterations, seed 0), resampling studies for internal B-mode data, patients for CAMUS and frames for internal Doppler and for MIMIC-IV-ECHO precision, with paired resampling for comparisons between models. Reader agreement was summarized by Cohen’s κ with the prevalence-adjusted κ. Intervals for classes with fewer than 15 annotated boxes are reported but not interpreted.

## Data availability

The internal echocardiograms and annotations from Mayo Clinic contain protected health information and cannot be shared publicly under the terms of the institutional review board approval; requests for access to de-identified data for non-commercial research can be directed to the corresponding author and require approval from Mayo Clinic and a data use agreement. The CAMUS dataset is publicly available from its providers (https://www.creatis.insa-lyon.fr/Challenge/camus/). MIMIC-IV-ECHO is available to credentialed users on PhysioNet (https://doi.org/10.13026/307c-mr50) under the PhysioNet Credentialed Health Data Use Agreement.

## Ethical Approval

This study was approved by the Institutional Review Board at Mayo Clinic (protocol: 22-010944). Informed consent was waived in accordance with institutional policies for minimal risk retrospective research.

## Acknowledgements

This work was supported by the National Heart, Lung, and Blood Institute of the National Institutes of Health under award K08HL175205 to C.-J.C., and by the Mayo Clinic Department of Cardiovascular Medicine CV Prospective Award and the Mayo Clinic MC-AIR Seedling Award. The content is solely the responsibility of the authors and does not necessarily represent the official views of the National Institutes of Health.

## Author contributions

C.-J.C. conceived and designed the study, performed the analyses, led the reader and external validation studies, supervised the project and acquired funding. E.M. developed the models, annotated data and contributed to the reader and external validation studies. N.P. developed the software. H.S., N.B.A., K.A., F.E.A., S.A.E., I.G.S. and J.F. curated and annotated the data. C.A., R.A., M.G., G.K., J.K.O. and I.B. provided resources and critically reviewed the manuscript. B.J.E. provided input on the study design and supervised the work. C.-J.C. and E.M. wrote the manuscript. All authors reviewed and approved the final manuscript.

## Competing interests

The authors declare no competing interests.

**1. Supplementary Information is provided as a separate file.**

## References

1. Lang, R. M. et al. Recommendations for cardiac chamber quantification by echocardiography in adults: an update from the American Society of Echocardiography and the European Association of Cardiovascular Imaging. J. Am. Soc. Echocardiogr. 28, 1–39.e14 (2015).

2. Madani, A., Arnaout, R., Mofrad, M. & Arnaout, R. Fast and accurate view classification of echocardiograms using deep learning. *npj Digit*. Med. 1, 6 (2018).

3. Ouyang, D. et al. Video-based AI for beat-to-beat assessment of cardiac function. Nature 580, 252– 256 (2020).

4. Zhang, J. et al. Fully automated echocardiogram interpretation in clinical practice: feasibility and diagnostic accuracy. Circulation 138, 1623–1635 (2018).

5. Christensen, M., Vukadinovic, M., Yuan, N. & Ouyang, D. Vision–language foundation model for echocardiogram interpretation. Nat. Med. 30, 1481–1488 (2024).

6. Holste, G. et al. Complete AI-enabled echocardiography interpretation with multitask deep learning. JAMA 334, 306–318 (2025).

7. Vukadinovic, M. et al. Comprehensive echocardiogram evaluation with view primed vision language AI. Nature 650, 970–977 (2026).

8. Huang, H. et al. EchoMLLM: incentivizing echocardiographic video understanding with keyframe grounding and report generation. In Findings of the Association for Computational Linguistics: ACL 2026 20053–20071 (Association for Computational Linguistics, 2026).

9. Selvaraju, R. R. et al. Grad-CAM: visual explanations from deep networks via gradient-based localization. In Proc. IEEE International Conference on Computer Vision 618–626 (IEEE, 2017).

10. Saporta, A. et al. Benchmarking saliency methods for chest X-ray interpretation. *Nat*. Mach. Intell. 4, 867–878 (2022).

11. Arun, N. et al. Assessing the trustworthiness of saliency maps for localizing abnormalities in medical imaging. *Radiol*. Artif. Intell. 3, e200267 (2021).

12. Redmon, J., Divvala, S., Girshick, R. & Farhadi, A. You only look once: unified, real-time object detection. In Proc. IEEE Conference on Computer Vision and Pattern Recognition 779–788 (IEEE, 2016).

13. Carion, N. et al. End-to-end object detection with transformers. In Computer Vision – ECCV 2020 213–229 (Springer, 2020).

14. Kirillov, A., et al. Segment anything. In Proc. IEEE/CVF International Conference on Computer Vision (IEEE, 2023); 10.1109/ICCV51070.2023.00371.

15. Liu, S. et al. Grounding DINO: marrying DINO with grounded pre-training for open-set object detection. In Computer Vision – ECCV 2024 ( Springer, 2024 ) ; 10.1007/978-3-031-72970-6_3.

16. Balasubramani, M. et al. Automated left ventricle segmentation in echocardiography using YOLO: a deep learning approach for enhanced cardiac function assessment. Electronics 13, 2587 (2024).

17. Gol Gungor, D., Rao, B., Wolverton, C. & Guracar, I. View classification and object detection in cardiac ultrasound to localize valves via deep learning. Preprint at https://arxiv.org/abs/2311.00068 (2023).

18. Qiao, S., et al. Automatic detection of cardiac chambers using an attention-based YOLOv4 framework from four-chamber view of fetal echocardiography. Preprint at https://arxiv.org/abs/2011.13096 (2020).

19. Komatsu, M. et al. Detection of cardiac structural abnormalities in fetal ultrasound videos using deep learning. Appl. Sci. 11, 371 (2021).

20. Lu, Y., Li, K., Pu, B., Tan, Y. & Zhu, N. A YOLOX-based deep instance segmentation neural network for cardiac anatomical structures in fetal ultrasound images. IEEE/ACM Trans. Comput. Biol. Bioinform. 21, 1007–1018 (2024).

21. Chen, S.-H. et al. Modified YOLOv4-DenseNet algorithm for detection of ventricular septal defects in ultrasound images. Int. J. Interact. Multimed. Artif. Intell. 6, 101–108 (2021).

22. Chen, S.-H. et al. Optimizing object detection algorithms for congenital heart diseases in echocardiography: exploring bounding box sizes and data augmentation techniques. Rev. Cardiovasc. Med. 25, 335 (2024).

23. Bai, S., et al. Qwen2.5-VL technical report. Preprint at https://arxiv.org/abs/2502.13923 (2025).

24. Sellergren, A., et al. MedGemma technical report. Preprint at https://arxiv.org/abs/2507.05201 (2025).

25. Huang, S., Hou, Y., Liu, L., Yu, X. & Shen, X. Real-time object detection meets DINOv3. Preprint at https://arxiv.org/abs/2509.20787 (2025).

26. Siméoni, O., et al. DINOv3. Preprint at https://arxiv.org/abs/2508.10104 (2025).

27. Lin, T.-Y. et al. Microsoft COCO: common objects in context. In Computer Vision – ECCV 2014 740–755 (Springer, 2014).

28. Gow, B. et al. MIMIC-IV-Echo: echocardiogram matched subset (version 1.0.1). PhysioNet 10.13026/307c-mr50 (2026).

29. Leclerc, S. et al. Deep learning for segmentation using an open large-scale dataset in 2D echocardiography. IEEE Trans. Med. Imaging 38, 2198–2210 (2019).

